# Clinical subgroup clustering analysis in a systemic lupus erythematosus cohort from Western Pennsylvania

**DOI:** 10.1101/2020.11.12.20230789

**Authors:** Patrick Coit, Lacy Ruffalo, Amr H Sawalha

## Abstract

**Objective:** Systemic lupus erythematosus (SLE) is a complex heterogenous autoimmune disease that can affect multiple organs. We performed clinical clustering analysis to describe a lupus cohort from the University of Pittsburgh Medical Center.

**Methods:** A total of 724 patients who met the ACR classification criteria for SLE were included in this study. Clustering was performed using the ACR classification criteria and the partitioning around medoid method. Correlation analysis was performed using the Spearman’s Rho test.

**Results:** Patients with SLE in our cohort identify 3 district clinical disease subsets. Patients in Cluster 1 were significantly more likely to develop renal and hematologic involvement, and had overrepresentation in African-American and male lupus patients. Clusters 2 and 3 identified a milder disease, with a significantly less likelihood of organ complications. Patients in Cluster 2 are characterized by malar rash and photosensitivity, while patients in Cluster 3 are characterized by oral ulcers which is present in ∼90% of patients within this cluster. The presence of photosensitivity or oral ulcers appears to be protective against the development of lupus nephritis in our cohort.

**Conclusions:** We describe a large cohort of SLE from Western Pennsylvania and identify 3 distinct clinical disease subgroups. Clustering analysis might help to better manage and predict disease complications in heterogenous diseases like lupus.

## Introduction

Systemic lupus erythematosus (SLE or lupus) is a chronic remitting-relapsing autoimmune disease characterized by the production of antinuclear antibodies. Lupus is heterogenous and can affect multiple organ systems ^1^. Although more commonly affects women, lupus tends to be more severe in men ^2^. In the United States lupus is more common and more severe in patients of African-American descent compared to European-American patients with the disease ^3^.

The etiology of lupus is not fully understood. Genetic and environmental factors are thought to be involved in the pathogenesis of lupus ^4^. Further, a clear role for epigenetic dysregulation in the pathogenesis lupus has been established ^5; 6^. The clinical heterogeneity of lupus is suggested to reflect variability in the underlying genetic background, epigenetic modifications, and immunologic dysregulation, among individual lupus patients ^7-10^. While lupus is unified by the presence of autoantibodies directed against self-nuclear antigens, clinical and molecular heterogeneity of the disease is an important factor hindering the success of clinical trials in lupus ^11^.

In this report, we describe a subset of lupus patients enrolled in the University of Pittsburgh Medical Center Lupus Cohort who meet the American College of Rheumatology (ACR) classification criteria for SLE ^12^. We implement a subgroup clinical clustering analysis and characterize 3 district clinical subsets of lupus in our Lupus Cohort.

## Methods

### Patients

We studied a subset of patients included in our UPMC Lupus Cohort who met the American College of Rheumatology classification criteria for SLE ^12^. All patients were evaluated in our clinics between January 2018 and April 2020. A total of 724 patients were studied.

### Clustering

The 11 ACR classification criteria for SLE were used as input for calculating a distance matrix using Gower’s Distance method using the *cluster* (v2.1.0) package in R. This method is intended for non-numeric data ^13^. All ACR criteria were entered as asymmetric binary values. Cluster group number (*k*) was determined *a priori* using the *NbClust* (v3.0) package ^14^. This method uses a collection of 30 clustering indices that suggested an optimal recommended *k* = 3. Clustering of Gower’s Distance matrix was performed using the partitioning around medoid (PAM) method in the *cluster* package that identifies clusters based around a single object with minimal dissimilarity to all objects with its cluster ^15^. PAM operates on the same principles as the *k*-means algorithm but is more robust to outliers ^16^. Assigned clusters had a combined average silhouette of 0.24 (Cluster 1 = 0.27, Cluster 2 = 0.25, and Cluster 3 = 0.19). Silhouette values can range from −1 to +1, with a higher value indicating a better cohesion of the objects within the cluster ^17^. Cluster assignments for each sample were used to test for differences in the distribution of sex, race/ethnicity, and presence of ACR criteria across clusters.

### Statistical analysis

A Pearson’s chi-square test was performed to compare sex and the presence of ACR criteria across clusters. A Fisher’s Exact test was performed to compare race/ethnicity across clusters. P values for the differences between the presence of the 11 ACR criteria across clusters were adjusted using the Benjamini-Hochberg method to account for multiple testing. Sex and race/ethnicity P values were reported unadjusted. Odds ratios and Fisher’s Exact test P values were calculated for sex and the presence of ACR criteria across clusters using the *epitools* (v0.5-10.1) package in R without correction for multiple testing ^18^. A significance threshold of P < 0.05 was used for all statistical testing. Correlation analysis was performed using the non-parametric Spearman’s Rho test with Benjamini-Hochberg FDR-adjusted P values reported to correct for multiple testing using the *correlations* (v0.4.0) package in R ^19^.

## Results

We evaluated a total of 724 lupus patients included in the Lupus Cohort at the University of Pittsburgh Medical Center. These patients represent a subset of our Lupus Cohort who meet the American College of Rheumatology classification criteria for SLE, and were evaluated at our center between January 2018 and March 2020.

Our study population included 672 female and 52 male lupus patients, and are 73% (n=529) European-American, 23% (n=168) African-American, 2% (n=16) Asians, and <2% (n=11) others (**Table 1**). The average and median age of our patients are 48 and 47 years, respectively (range 19 to 86).

**Table 1:** Clinical characteristics of 3 subgroups of patients with lupus in our cohort

|  | **All patients**<br>**(n= 724)** | **Cluster 1**<br>**(n= 270)** | **Cluster 2**<br>**(n= 179)** | **Cluster 3**<br>**(n= 275)** | **P value** |
| --- | --- | --- | --- | --- | --- |
| **Race/ethnicity** |  |  |  |  |  |
| White | 529 (73%) | 157 (58%) | 146 (82%) | 226 (82%) | 2.04E-09 |
| Black | 168 (23%) | 98 (37%) | 29 (16%) | 41 (15%) | - |
| Asian | 16 (2%) | 9 (3%) | 3 (2%) | 4 (1%) | - |
| Other/Not Reported | 11 (<2%) | 6 (2%) | 1 (<1%) | 4 (1%) |  |
| **Sex** |  |  |  |  |  |
| Female | 672 (93%) | 242 (90%) | 166 (93%) | 264 (96%) | 0.0158 |
| Male | 52 (7%) | 28 (10%) | 13 (7%) | 11 (4%) | - |
| **Manifestations** |  |  |  |  |  |
| Malar rash | 218 (30%) | 39 (14%) | 125 (70%) | 54 (20%) | 1.77E-39 |
| Discoid rash | 95 (13%) | 48 (18%) | 18 (10%) | 29 (11%) | 0.0201 |
| Photosensitivity | 353 (49%) | 29 (11%) | 142 (79%) | 182 (66%) | 7.11E-56 |
| Oral ulcers | 322 (44%) | 39 (14%) | 37 (21%) | 246 (89%) | 6.14E-79 |
| Arthritis | 623 (86%) | 218 (81%) | 163 (91%) | 242 (88%) | 0.005744 |
| Serositis | 232 (32%) | 91 (34%) | 49 (27%) | 92 (33%) | 0.334 |
| Renal disorder | 119 (16%) | 82 (30%) | 19 (11%) | 18 (7%) | 5.79E-14 |
| Neurologic disorder | 30 (4%) | 14 (5%) | 5 (3%) | 11 (4%) | 0.455 |
| Hematologic disorder | 277 (38%) | 204 (76%) | 19 (11%) | 54 (20%) | 7.11E-56 |
| Immunologic disorder | 496 (69%) | 250 (93%) | 147 (82%) | 99 (36%) | 1.22E-48 |
| Positive ANA | 674 (93%) | 262 (97%) | 160 (89%) | 252 (92%) | 0.00562 |

To further characterize the patterns of disease involvement in our lupus patients, we performed a medoid clustering analysis using the 11 ACR classification criteria for lupus. The analysis revealed that our lupus patients cluster in 3 distinct clinical clusters (**Figure 1**).

**Figure 1.**
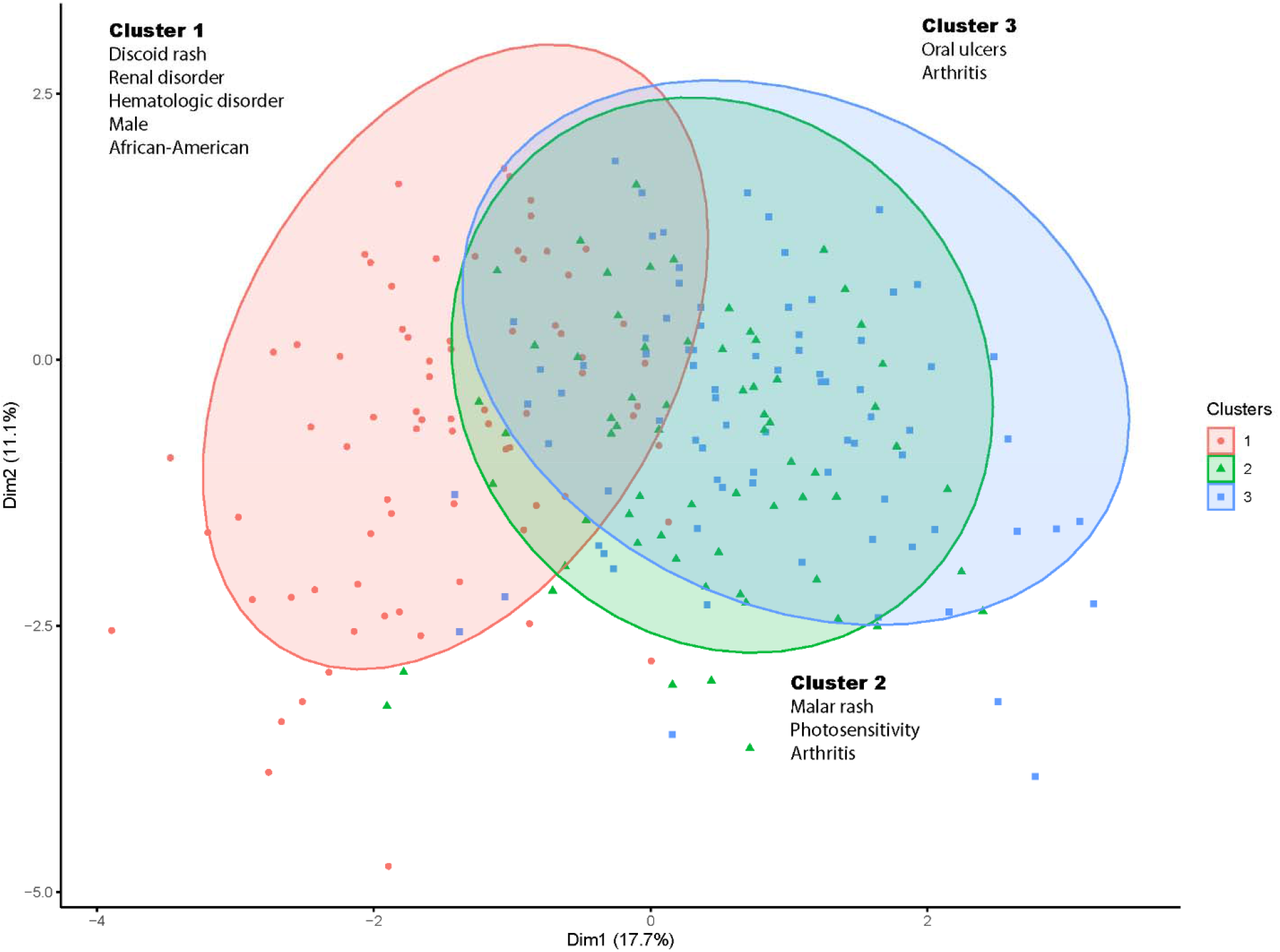
Clustering analysis of 724 lupus patients reveals 3 disease subsets. Clusters were determined using portioning around medoids method applied to a Gower’s Distance matrix of 11 ACR criteria reported for all patients.

Lupus Cluster 1 includes 270 (37%) patients with overrepresentation of organ specific manifestations. This includes renal involvement in 30% of patients, compared to 11% and 7% in Clusters 2 and 3, respectively (P=5.79E-14), hematologic involvement (76%, compared to 11% and 20% in Clusters 2 and 3, respectively, P= 7.11E-56), and discoid rash (18%, compared to 10% and 11% in Clusters 2 and 3, respectively, P= 0.02). As shown in **Table 1**, among all of lupus patients in our cohort that have renal involvement (n=119), hematological involvement (n=277), and discoid rash (n=95), 69%, 74%, 51%, respectively, are in Cluster 1. Not unexpectedly, the majority of our African-American lupus patients (98 of 168 patients) were in this cluster, which is also enriched with our male lupus patients (28 of 52 male patients in our cohort) (**Table 1**).

Patients in Cluster 2 (25%, n=179) were more likely to have non-chronic cutaneous involvement including malar rash (70% of patients, P=1.77E-39) and photosensitivity (79% of patients, P= 7.11E-56), and arthritis (91% of patients, P= 0.0057). Cluster 3 (38%, n=275) is characterized by oral ulcers in the vast majority of patients (89%, n= 246) and has the lowest rate of renal involvement among all 3 clusters (7%) (**Table 1**).

We next determined the odds of developing specific lupus features for patients in any given cluster (**Table 2**). Patients in Cluster 1 were 3.7 and 6.25 times more likely to develop lupus renal involvement compared to Clusters 2 and 3, respectively (P= 5.16E-07 and 2.59E-13), and 25 and 12.5 times more likely to have hematologic involvement (P= 6.25E-45 and 5.63E-41). Cluster 2 patients were 13.6 and 31.5 times more likely to have malar rash and photosensitivity, respectively, compared to Cluster 1 (P= 1.85E-33 and 1.69E-51). Meanwhile, patients in Cluster 3 were ∼50 times more likely to have oral ulcers (OR= 49.54, P= 1.20E-76) and were protected from lupus nephritis (OR= 0.16, P= 2.59E-13) compared to patients in Cluster 1 (**Table 2**)

**Table 2.** Odds ratios for differences in clinical characteristics and manifestations between the lupus subgroups identified in our study. Odds ratio values and 95% confidence intervals (CI) in Clusters 2 and 3 versus Cluster 1 are depicted.

| Variable | Cluster | Odds ratio (vs. Cluster 1) | Lower 95%CI | Upper 95%CI | P value |
| --- | --- | --- | --- | --- | --- |
| Sex (Female) | Cluster 2 | 1.48 | 0.72 | 3.20 | 3.17E-01 |
|  | Cluster 3 | 2.77 | 1.30 | 6.31 | 4.43E-03 |
| Malar rash | Cluster 2 | 13.60 | 8.40 | 22.48 | 1.85E-33 |
|  | Cluster 3 | 1.45 | 0.90 | 2.34 | 1.12E-01 |
| Discoid rash | Cluster 2 | 0.52 | 0.27 | 0.95 | 2.89E-02 |
|  | Cluster 3 | 0.55 | 0.32 | 0.92 | 1.91E-02 |
| Photosensitivity | Cluster 2 | 31.50 | 18.23 | 56.06 | 1.69E-51 |
|  | Cluster 3 | 16.16 | 10.08 | 26.61 | 3.60E-43 |
| Oral ulcers | Cluster 2 | 1.54 | 0.91 | 2.61 | 9.52E-02 |
|  | Cluster 3 | 49.54 | 29.23 | 87.04 | 1.20E-76 |
| Arthritis | Cluster 2 | 2.43 | 1.31 | 4.72 | 2.94E-03 |
|  | Cluster 3 | 1.75 | 1.06 | 2.90 | 2.45E-02 |
| Serositis | Cluster 2 | 0.74 | 0.48 | 1.14 | 1.77E-01 |
|  | Cluster 3 | 0.99 | 0.68 | 1.43 | 1.00E+00 |
| Renal disorder | Cluster 2 | 0.27 | 0.15 | 0.48 | 5.16E-07 |
|  | Cluster 3 | 0.16 | 0.09 | 0.28 | 2.59E-13 |
| Neurologic disorder | Cluster 2 | 0.53 | 0.15 | 1.51 | 2.42E-01 |
|  | Cluster 3 | 0.76 | 0.31 | 1.85 | 5.45E-01 |
| Hematologic disorder | Cluster 2 | 0.04 | 0.02 | 0.07 | 6.25E-45 |
|  | Cluster 3 | 0.08 | 0.05 | 0.12 | 5.63E-41 |
| Immunologic disorder | Cluster 2 | 0.37 | 0.19 | 0.69 | 8.72E-04 |
|  | Cluster 3 | 0.05 | 0.03 | 0.08 | 3.99E-47 |
| Positive ANA | Cluster 2 | 0.26 | 0.10 | 0.63 | 1.78E-03 |
|  | Cluster 3 | 0.34 | 0.13 | 0.79 | 8.70E-03 |

A correlation analysis between the 11 ACR criteria was performed in our lupus patients. We detected a significant positive correlation between fulfilling the immunologic disorder criterion and both renal involvement and hematologic disorder (P<0.001 and <0.01, respectively). The presence of either photosensitivity or oral ulcers in our lupus patients was negatively correlated with the presence of renal disorder, hematologic disorder, and immunologic disorder (P<0.001 for all correlations) (**Figure 2**).

**Figure 2:**
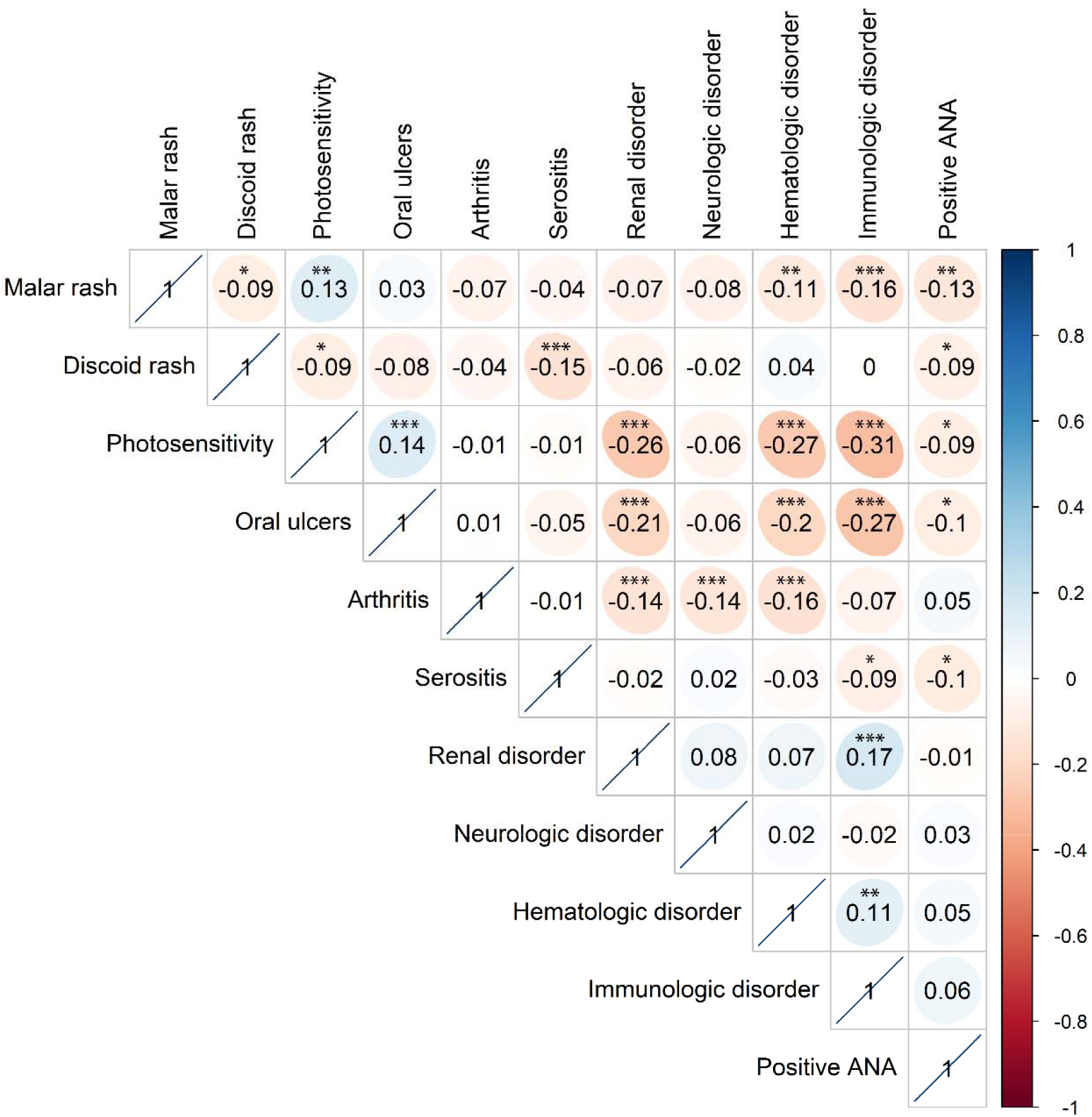
Correlation matrix of 11 ACR criteria reported for 724 lupus patients included in our study. Correlation values were calculated using Spearman’s Rho test. P values were adjusted for multiple testing using the Benjamini-Hochberg false discovery rate method and adjusted P values are reported. *, P < 0.05; **, P < 0.01; ***, P < 0.001.

## Discussion

Systemic lupus erythematosus is a heterogenous remitting-relapsing chronic autoimmune disease. In this report, we describe a lupus cohort from a single tertiary referral center in Western Pennsylvania. Clustering analysis based on the ACR classification criteria for systemic lupus erythematosus identified 3 distinct clinical lupus clusters. 37% of our lupus patients are within a cluster of a more severe disease characterized by renal and hematologic involvement, 25% are in a cluster characterized by malar rash and photosensitivity, and the remaining 38% are in a cluster characterized by the presence of oral ulcers. Patients in the latter two clusters have less severe lupus with a significantly lower frequency of organ complications such as renal involvement. Intriguingly, our data suggest that the presence of photosensitivity or oral ulcers in lupus patients is protective against the development of lupus nephritis.

Clinical clustering in heterogenous diseases helps to identify disease subsets and might have value in predicting patterns of disease involvement and expected disease severity and organ complications ^20^. In lupus, our data suggest 3 clinical disease subsets with distinct patterns of clinical manifestations and differences in the odds of developing organ involvement. These data might have implications in the management of lupus patients.

Whether difference in the molecular mechanisms underlying lupus influence or determine the clinical clustering we observed in our patients remains to be determined. If that were to be the case, then perhaps clinical clustering might be a useful tool to reduce disease heterogeneity in lupus clinical trials with the premise that this might improve the likelihood of achieving a successful outcomes in lupus trials ^21^.

Our results are derived from a single cohort of lupus patients, and might not necessarily reflect the clinical subsets of lupus in other lupus cohorts from different geographic location or different ancestral groups of patients. Expanding these observations and examining clinical clustering in lupus patients from across different ancestries and locations are certainly warranted.

In summary, we describe lupus patients from our Lupus Cohort at the University of Pittsburgh Medical Center and identify distinct clinical subsets of lupus characterized by a specific pattern of disease and organ involvement. These data might have implication in the clinical care of lupus patients. Further, clinical clustering might be a useful tool to reduce disease heterogeneity and improve outcomes in clinical trials in lupus and similar complex autoimmune diseases.

## Data Availability

All data are provided in the manuscript

## Acknowledgements

This work was supported by the National Institute of Allergy and Infectious Diseases of the National Institutes of Health grant number R01AI097134, and the Lupus Research Alliance.

